# Leveraging next-generation phenotyping for ACMG classification from VUS to likely pathogenic in Mowat-Wilson syndrome

**DOI:** 10.1101/2025.02.12.25321927

**Authors:** Tzung-Chien Hsieh, Dylan Todd, Taylor Warner, Kayla Blankenship, Dimah Saade, Hannah Weiland, Meghna Ahuja Bhasin, Hannah Klinkhammer, Jing-Mei Li, Peter M. Krawitz, Wei-Liang Chen, Ho-Ming Luk, Moon Ley Tung, Bharatendu Chandra

## Abstract

**Background:** Next-generation phenotyping (NGP) tools, such as GestaltMatcher, have revolutionized the diagnosis of rare genetic disorders through computational facial analysis. While NGP has been widely integrated into differential diagnosis workflows, its application in variant reclassification within the ACMG framework remains underexplored.

**Methods:** We applied GestaltMatcher to a pediatrics patient with a severe neurodevelopmental disorder, suspected Mowat-Wilson syndrome (MWS), and a de novo *ZEB2* variant initially classified as a variant of uncertain significance (VUS). In addition to facial image analysis, we utilized the PEDIA framework, integrating Human Phenotype Ontology (HPO) terms and simulated exome data to refine variant prioritization. Bayesian likelihood modeling was used to establish Gestalt score thresholds for PP4 evidence levels (supporting, moderate, strong, and very strong). Brain MRI analysis was also performed to assess structural abnormalities characteristic of MWS.

**Results:** GestaltMatcher ranked MWS as the top differential diagnosis, and PEDIA integration further confirmed *ZEB2* as the most likely disease-causing gene. Three of the patient’s four facial images met the PP4 moderate threshold, while one met PP4 supporting. Based on these findings, the *ZEB2* variant was reclassified as Likely Pathogenic. MRI analysis revealed subtle corpus callosum thinning, consistent with MWS. Additionally, a validation case of an infant with molecularly confirmed MWS demonstrated the capability of GestaltMatcher to prioritize the diagnosis solely based on infant facial features.

**Conclusion:** This study highlights the potential of NGP-driven facial phenotyping and multimodal integration in variant reclassification. The results support the broader application of AI-assisted phenotyping to improve diagnostic accuracy and ACMG-based variant interpretation, particularly in neurodevelopmental disorders with distinct facial features.

## Introduction

Next-generation phenotyping (NGP) approaches, such as GestaltMatcher^1^, have been widely adopted for computational facial image analysis in the diagnosis of rare disorders. Over recent years, GestaltMatcher has not only provided a ranked list of differential diagnoses but also contributed to variant prioritization through the PEDIA framework^2^. The German National Rare Disorder Project, TRANSLATE-NAMSE^3^, demonstrated that integrating Human Phenotype Ontology (HPO)^4^ analysis via CADA^5^, facial image analysis through GestaltMatcher, and CADD scores^6^ derived from exome data significantly improved diagnostic accuracy. While numerous studies have highlighted the effectiveness of NGP in diagnosing rare disorders^7–9^, the further integration of GestaltMatcher into the ACMG classification framework^10^, using Bayesian approaches^11^, remains an area for exploration.

In this study, we sought to test the utility of GestaltMatcher in a patient with severe neurodevelopmental disorder and suspected Mowat-Wilson syndrome (MIM:235730), harboring a *ZEB2* variant initially classified as a variant of uncertain significance (VUS). Using both clinical phenotyping and GestaltMatcher to analyze the patient’s frontal image, we reclassified the variant as likely pathogenic. Additionally, we expanded the use of NGP to include brain MRI analysis, identifying structural brain abnormalities unique to patients with Mowat-Wilson syndrome compared to healthy controls. This demonstrated the potential of NGP technology to integrate data from multiple modalities, laying the groundwork for multimodal learning applications in medical imaging and diagnosis.

## Materials and Methods

### Patient

The proband with a severe neurodevelopmental disorder was evaluated at the Neurology and Genetics clinic at the University of Iowa Hospitals and clinics. Informed consent was obtained for this study through the UI rare diseases biorepository. We obtained photography and bio repository consents as well as whole exome sequencing raw data from Variantyx in compliance with the authors institutional policies.

### Facial phenotyping with GestaltMatcher

GestaltMatcher is an NGP approach that trains on a patient’s two-dimensional frontal images to learn the facial phenotype of the disease. In this study, we employed GestaltMatcher-Arc^12^, an advanced version of GestaltMatcher, which integrates model ensembles and test-time augmentation to enhance performance. For simplicity, we will refer to GestaltMatcher-Arc as GestaltMatcher throughout the following sections. GestaltMatcher was trained on a dataset of 9,671 images from 7,294 patients diagnosed with 275 distinct disorders, all sourced from the GestaltMatcher Database (GMDB)^13^. The models encoded each image into 12 Facial Phenotype Descriptors (FPD). The facial phenotypic similarity between two patients is quantified by calculating the average cosine distance across the 12 pairs of FPDs derived from their images. A smaller cosine distance indicates higher similarity, as it suggests the two patients are located closer together within the Clinical Face Phenotype Space (CFPS). To further simplify interpretation, we transformed the cosine distance into a Gestalt score, defined as 1.3−cosine distance. This transformation ensures that a higher Gestalt score corresponds directly to a greater degree of similarity, making it more intuitive for analysis. Finally, the Gestalt scores for each disorder can be ranked, enabling the model to generate a prioritized list of differential diagnoses.

### Calculating the thresholds for gestalt score to PP4 criteria

For each image, GestaltMatcher quantifies the Gestalt score for every syndrome present in GMDB. We then used these scores to determine thresholds for PP4_SUP (supporting), PP4_MOD (moderate), PP4_STR (strong), and PP4_VSTR (very strong), following the Bayesian framework proposed by Tavtigian et al. ^11^. In GMDB, 75 images from 51 patients with Mowat-Wilson syndrome were included. 66 of these images were included during GestaltMatcher model training. To establish the PP4 thresholds, we divided the data into a training set (61 images) and a validation set (5 images). The remaining nine images were not used in model training and were reserved for testing. The training set comprised 61 Mowat-Wilson syndrome images and 610 control images representing other syndromes. The validation set included 5 Mowat-Wilson syndrome images alongside 50 control images. The PP4 criteria at each evidence level were defined using likelihood ratios: supporting (2.08), moderate (4.3), strong (18.7), and very strong (350). We computed the likelihood ratios for different Gestalt score thresholds using the training set and subsequently validated them on the validation set.

### Prioritization of exome data by integrating facial image and HPO analysis (PEDIA)

The PEDIA approach integrates Human Phenotype Ontology (HPO) terms, facial image analysis, and exome data to prioritize disease-causing variants. For HPO-based phenotypic similarity, we utilized the CADA score, while facial image analysis was performed using the GestaltMatcher score. To map these scores at the gene level, we linked each disorder to its associated gene, ensuring a per-gene score representation. Since original exome data in VCF format was unavailable, we simulated exome data by introducing disease-causing mutations into healthy controls selected from the 1000 Genomes Project.

For variant pathogenicity score, we annotated each variant using CADD version 1.6 and assigned the highest CADD score per gene. These three scores—CADA, GestaltMatcher, and CADD—were then combined using a Support Vector Machine (SVM) model. The model was trained on 679 patient cases with 105 different disorders, where the three scores served as input features, and the output was a ranked list of candidate genes, each assigned a PEDIA score. To evaluate performance, we ranked genes in descending order of PEDIA scores. We assessed accuracy using top-k ranking, which measures whether the disease-causing gene appears within the top-k positions.

### Brain MRI analysis

A 3-Tesla MRI Brain was performed for detailed clinical phenotyping for the neurodevelopmental disorder in this patient. The MRI sequences used for the analysis included T1 and T2 weighted, Fluid attenuation inversion recovery (FLAIR) images with axial, coronal, and sagittal sections.

## Results

### Patient clinical and molecular analysis

The Proband is a female who first presented with concerns regarding delayed developmental milestones at year of 1-5. Her scores on the appropriate ASQ-3 at year of 1-5 were within the reference range across all developmental domains. Behavioral observations did not raise concerns about autism spectrum disorder. Despite these early reassuring findings, proband’s parents continue to express concerns about her developmental progress. At year of age 1-5, her gross motor development was notably delayed with rudimentary crawling. The proband, subsequently received a detailed developmental evaluation where she was found to have limited expressive language with intermittent simple words such as “mama,” “that,” and unintelligible vocalizations. ASQ-3 revealed significant delays in communication, gross motor, fine motor, and personal-social domains. The MCHAT-R was also administered and placed her in the medium-risk category for ASD. Due to this finding, the STAT was performed and indicated that the proband did not meet criteria autism diagnosis. The psychologist performing her assessment noted that the proband’s interactive behaviors, use of gestures, and joint attention were her strengths.

A sedated brain MRI was performed at year of age 1-5, which was interpreted as possible subtle thinning of the corpus callosum body without other structural abnormalities. At year of age 1-5, the proband underwent a comprehensive evaluation by our institutional developmental pediatric center. The interdisciplinary assessment confirmed the diagnosis of global developmental delay. Despite her delays, the proband exhibited functional pre-academic skills comparable to peers with intervention services.

She was diagnosed with nonaccommodative bilateral esotropia at year of age 1-5 and underwent bilateral medial rectus recession to address this condition. Some residual minor esotropia has responded well to patching and glasses.

At year of age 1-5, proband was evaluated by a neuromuscular specialist due to her motor delay. Examination revealed no evidence of muscle weakness or facial weakness with normal reflexes. She was found to have sialorrhea that fluctuates with her energy level. She was found to have a normal serum creatine kinase. A primary neuromuscular etiology for her developmental delays was considered unlikely, and she was referred to genetic counseling to investigate the etiology of global developmental delay.

Initial diagnostic workup including a Chromosomal microarray (CMA) and CK testing were unremarkable. A clinical Whole exome sequencing (WES) identified a de novo variant of uncertain significance in the ZEB2 gene (c.3237del; p.Tyr1080ThrfsTer28). This variant introduces a premature termination codon in exon 10 which was predicted to produce a truncated protein rather than undergo nonsense-mediated mRNA decay. Loss of function is a known pathogenic mechanism for Mowat-Wilson syndrome (MWS), and variants in exon 10 have been reported in affected individuals.

Family history: A three-generation pedigree was obtained by the authors. Consanguinity was denied. One parent’s ancestry is of unknown descent, while the other parent’s ancestry is of Hispanic descent. No Ashkenazi Jewish ancestry was reported. There was no history of genetic diagnoses, neurologic diseases, recurrent miscarriages, birth defects, cognitive or physical disabilities within the family. Proband has a full sibling with history of hypospadias. She also had a full younger sibling who died at year of age 1-5 from sudden infant death syndrome (SIDS).

Proband was seen by the authors in a medical genetics clinic at year of age 1-5. She was noted to have a happy disposition which can be characteristic of children with Mowat-Wilson syndrome. Her dysmorphic facial features include a broad forehead, thickened eyebrows, and a rounded nasal tip with a high nasal bridge, and short columella **(Figure 1)**. Her ears are low-set and posteriorly rotated with upturned lobes, and she exhibits a full bottom lip and a pointed chin **(Figure 1)**. Her facies had a hypotonic impression. Ophthalmologic findings include left eye esotropia, for which she is wearing corrective strabismus glasses. Limb examination reveals clinodactyly of the third, fourth, and fifth digits of the lower extremities, as well as widened lower extremity digits **(Figure 1)**. We then used GestaltMatcher for external validation of our clinical phenotyping. GestaltMatcher predicted Mowat-Wilson syndrome as the top priority genetic disorder based on facial gestalt. With this information, we were able to reclassify the *ZEB2* variant as pathogenic (**Table 1**) and recommended echocardiogram and renal ultrasound for disease-specific surveillance related to Mowat-Wilson syndrome, given the high prevalence of cardiac, renal, and urinary tract anomalies in this condition.

**Table 1.** Proband’s variant classification based on ACMG classification and proposed NGP-based PP4 criteria.

| Variant | Zygoty | Inheritance | Initial ACMG Classification of Variant | NGP-based ACMG classification |
| --- | --- | --- | --- | --- |
| **ZEB2(NM_014795.4)**<br>**c.3237del, p.**<br>**Tyr1080ThrfsTer28** | Heterozygous | Denovo | VUS<br>PVS1_Moderate,<br>PS2_Moderate,<br>PM2_Supporting | Likely pathogenic<br>PVS1_Very Strong,<br>PS2_Strong<br>PM2_Moderate,<br>PP4_Moderate |

**Table 2.**
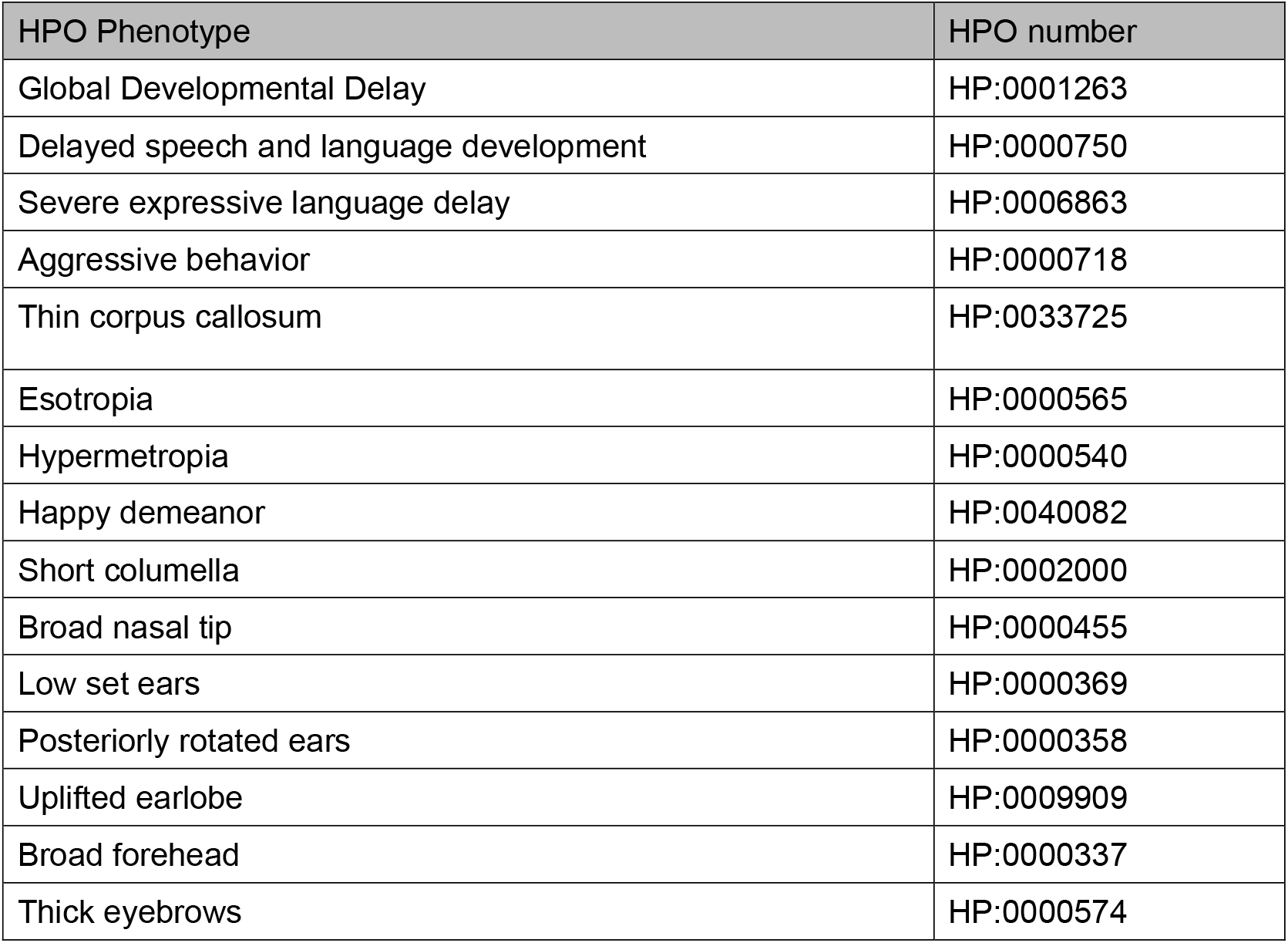
Proband’s clinical description in HPO terms.

| HPO Phenotype | HPO number |
| --- | --- |
| Global Developmental Delay | HP:0001263 |
| Delayed speech and language development | HP:0000750 |
| Severe expressive language delay | HP:0006863 |
| Aggressive behavior | HP:0000718 |
| Thin corpus callosum | HP:0033725 |
| Esotropia | HP:0000565 |
| Hypermetropia | HP:0000540 |
| Happy demeanor | HP:0040082 |
| Short columella | HP:0002000 |
| Broad nasal tip | HP:0000455 |
| Low set ears | HP:0000369 |
| Posteriorly rotated ears | HP:0000358 |
| Uplifted earlobe | HP:0009909 |
| Broad forehead | HP:0000337 |
| Thick eyebrows | HP:0000574 |

**Figure 1.**
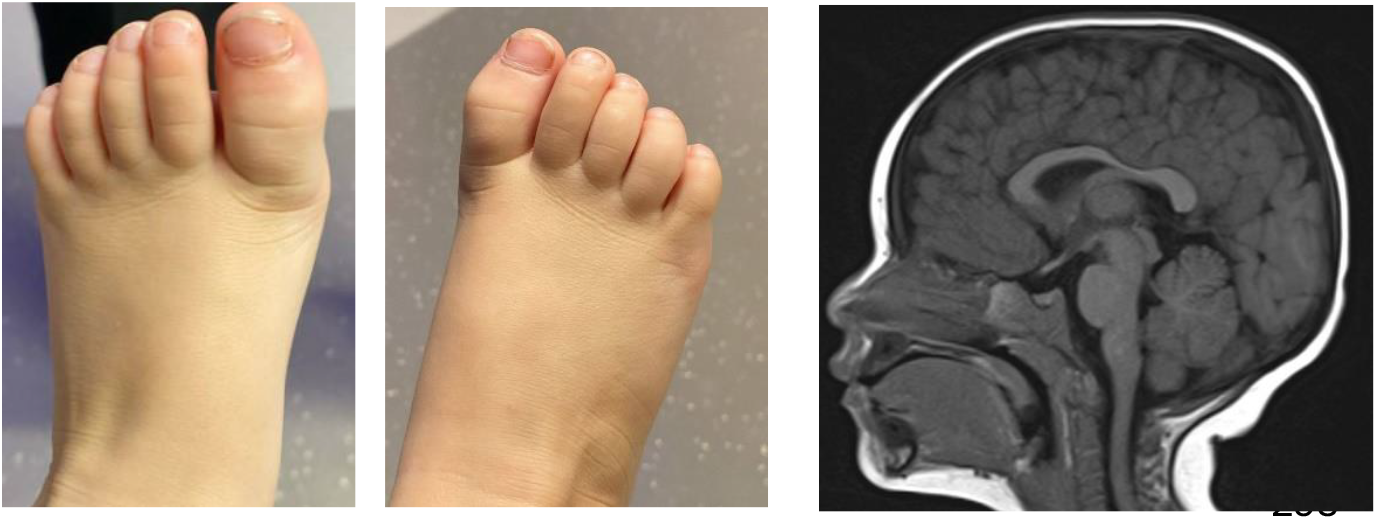
Facial Profile, feet, and T1Sag MRI Brain of the Proband. Facial images are removed due to regulation of medRxiv.

### Brain MRI findings

Thinning of the body of corpus callosum was the most distinct finding noted on analysis. There were no white matter changes abnormalities, ventriculomegaly, cortical dysplasia, basal ganglia or cerebellar abnormalities noted. Interestingly, the corpus callosum abnormality was in keeping with the findings of a large study that compared neuroimaging findings in Mowat-Wilson syndrome^14^.

### Facial image analysis facilitates the ACMG reclassification

We further applied the PEDIA approach to the four available photos, HPO terms, and simulated exome data containing the spiked-in disease-causing mutation. The results are summarized in **Table 3**. Using the molecular deleterious score (CADD score), the disease-causing mutation ranked fifth. Based on the HPO-derived scoring (CADA score), it achieved the top rank. Similarly, GestaltMatcher, which evaluates facial features, also ranked the disease-causing mutation first. When combining all three scores into the PEDIA score, the mutation achieved the top rank.

**Table 3.** Results of PEDIA, CADA, CADD, and GestaltMatcher scores for four different photos. The HPO (CADA score) and exome data (CADD score) remain constant across all images, with only the GestaltMatcher and PEDIA scores varying. Photos 1 and 2 are reported in **Figure 1** and photos 3 and 4 are not shown.

|  | CADA |  | CADD |  | GestaltMatcher |  | PEDIA |  |
| --- | --- | --- | --- | --- | --- | --- | --- | --- |
| Photo | Score | Rank | Score | Rank | Score | Rank | Score | Rank |
| 1 | 0.235 | 1 | 33.0 | 5 | 0.593 | 1 | 0.57 | 1 |
| 2 | 0.235 | 1 | 33.0 | 5 | 0.58 | 1 | 0.48 | 1 |
| 3 | 0.235 | 1 | 33.0 | 5 | 0.647 | 1 | 0.91 | 1 |
| 4 | 0.235 | 1 | 33.0 | 5 | 0.547 | 1 | 0.27 | 1 |

Since the mutation was initially classified as a VUS, we applied the proposed NGP-based Gestalt score to assess its evidence level under the PP4 criteria. First, we calculated the distribution of Gestalt scores for images of Mowat-Wilson syndrome patients and control images. As shown in **Figure 2**, the Gestalt scores effectively distinguish between Mowat-Wilson syndrome patients and controls, demonstrating the capability of facial image analysis in identifying the syndrome.

**Figure 2.**
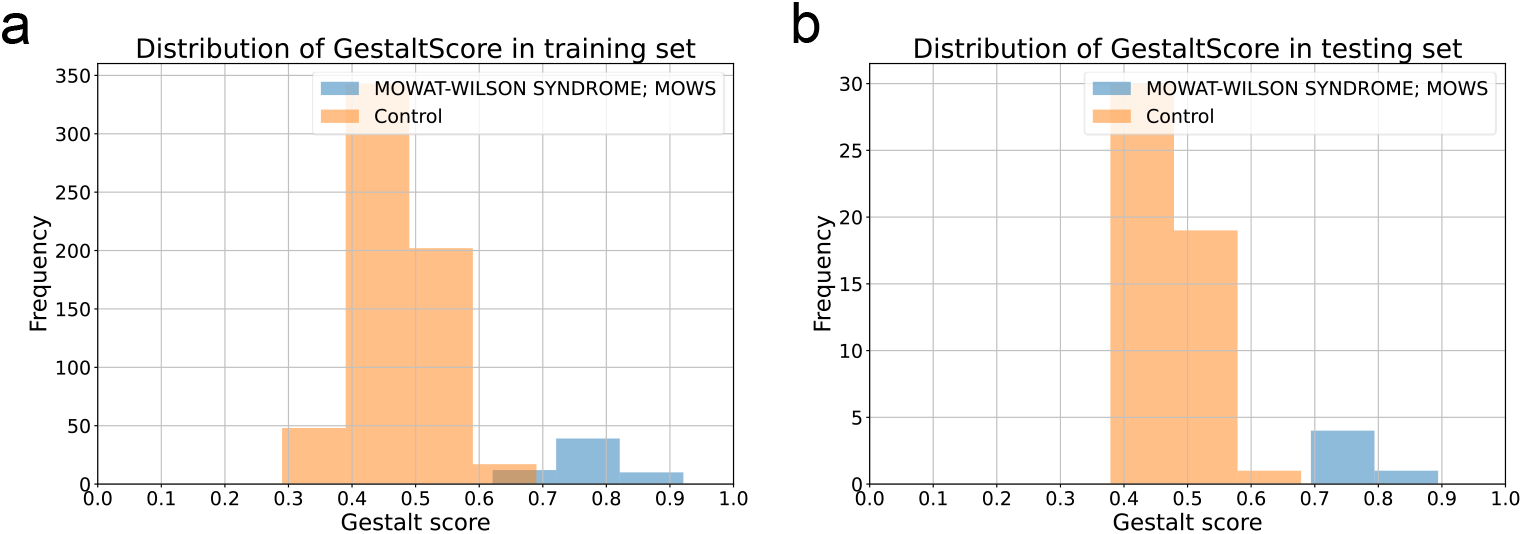
Distribution of Gestalt scores between patients with Mowat-Wilson syndrome and controls for training and testing datasets.

Next, we computed the likelihood ratios corresponding to different Gestalt scores, as reported in **Figure 3**. Based on these values, we established threshold Gestalt scores for each PP4 evidence level: supporting (0.512), moderate (0.5712), strong (0.665), and very strong (0.7671). The corresponding likelihood ratios, sensitivities, and specificities are presented in **Table 4**. Among the four analyzed images, three met the PP4 moderate threshold, while one met the PP4 supporting threshold. Consequently, all four facial images contributed to reclassifying the VUS as Likely Pathogenic (**Table 1**).

**Table 4.** Thresholds for each evidence level, along with the corresponding Likelihood Ratio (LR), Sensitivity, and Specificity for both training and testing datasets.

| Evidence level | Threshold | Likelihood ratio (LR) |  |  | Sensitivity |  | Specificity |  |
| --- | --- | --- | --- | --- | --- | --- | --- | --- |
|  |  | ACMG | Training | Testing | Training | Testing | Training | Testing |
| very strong | 0.7671 | 350 | inf | inf | 0.5574 | 0.4 | 1 | 1 |
| strong | 0.665 | 18.7 | 570.00 | inf | 0.9016 | 1 | 0.99 | 1 |
| moderate | 0.5712 | 4.3 | 19.06 | 24.99 | 1 | 1 | 0.94 | 0.96 |
| supporting | 0.512 | 2.08 | 4.32 | 3.12 | 1 | 1 | 0.76 | 0.76 |

**Figure 3.**
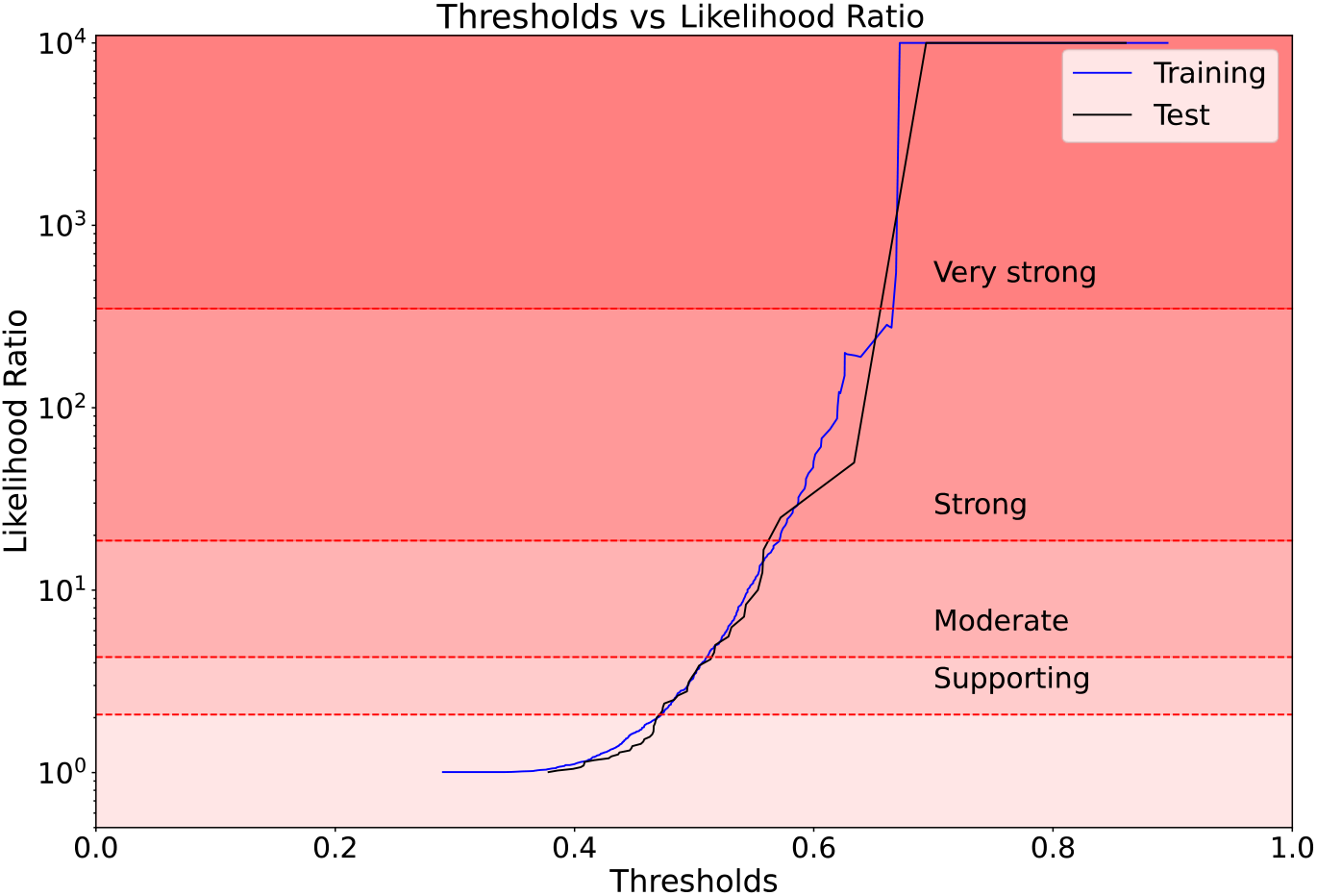
Distribution of Likelihood Ratios with corresponding thresholds. The Likelihood Ratios for supporting, moderate, strong, and very strong evidence levels are defined as 2.08, 4.3, 18.7, and 350, respectively.

### Validation of NGP using another case of MWS from our internal cohort

To perform validation of dysmorphology evaluation of model internally at our institution, we analysed facial profile of a male infant with molecularly confirmed diagnosis of MWS due to a pathogenic variant in *ZEB2* (c.1489C>T, p.Q497*). We tested GestaltMatcher agnostic classification by analysing only the facial profile of this case without entering additional clinical phenotype details. Notably, GestaltMatcher was able to prioritise MWS as the top ranked condition even with the facial profile during infancy. This is significant because MWS dysmorphology features are not that apparent during infancy, and it suggests that this tool could possibly be used to identify infants with genetic forms of neurodevelopmental disorder with a facial image alone.

## Discussion

Neurodevelopmental disorders affect over 3% of the population, often presenting with a highly heterogeneous clinical phenotype, and often result in impairment of cognitive, social, and motor developmental milestones in children. Rapid advances in next-generation sequencing technologies have played a crucial role in improving diagnostic yield for complex neurodevelopmental disorders. Over the past decade, several genes have been identified to be associated with monogenic forms of syndromic or non-syndromic neurodevelopmental disorders. In fact, the clinical adoption of exome or genome sequencing has resulted in rapid molecular diagnosis in a significant proportion of Neurodevelopmental disorders. However, VUS is a major challenge in next-generation sequencing approaches. Variant prioritization is heavily dependent on phenotyping and given the limited access to clinical genetics service for most patients with suspected rare diseases, implementing deep phenotyping with high-quality dysmorphology exams is not feasible. Therefore, NGP could fill this gap as a bridge between non-geneticist providers and geneticists that is able to perform high-quality phenotyping based on facial features and other imaging data, where available.

In this study, we applied NGP-based analysis to a proband suspected of having Mowat-Wilson syndrome with a VUS in *ZEB2*. Using a GestaltMatcher-based Gestalt score, we objectively quantified the PP4 criteria and successfully reclassified the variant as LP. Facial analysis using GestaltMatcher accurately predicted Mowat-Wilson syndrome in both probands analyzed, reinforcing its diagnostic reliability. Furthermore, integrating phenotypic and molecular data within the PEDIA framework enhanced variant prioritization, demonstrating improved accuracy and efficiency.

An additional strength of this approach is its implementation as an on-premise solution^15^, allowing it to be run locally in clinical settings without requiring external data transfer. This ensures data security, facilitates reanalysis, and reduces reliance on commercial entities. Our findings provide a proof-of-concept for integrating NGP tools into variant classification workflows, potentially benefiting undiagnosed disease networks.

## Data Availability

All data produced in the present study are available upon reasonable request to the authors.

## Ethics statement and consent for publication

This case report adheres to the Helsinki Declaration standards as well as national guidelines on the ethical integrity of reporting case reports. A written informed consent for publication of any potentially identifiable images or data was obtained from the parent of the patient.

## References

1. Hsieh, T.-C. et al. GestaltMatcher facilitates rare disease matching using facial phenotype descriptors. Nat. Genet. 54, 349–357 (2022).

2. Hsieh, T.-C. et al. PEDIA: prioritization of exome data by image analysis. Genet. Med. 21, 2807–2814 (2019).

3. Schmidt, A. et al. Next-generation phenotyping integrated in a national framework for patients with ultrarare disorders improves genetic diagnostics and yields new molecular findings. Nat. Genet. 1–10 (2024).

4. Köhler, S. et al. The Human Phenotype Ontology in 2021. Nucleic Acids Res. 49, D1207–D1217 (2021).

5. Peng, C. et al. CADA: phenotype-driven gene prioritization based on a case-enriched knowledge graph. NAR Genom Bioinform 3, lqab078 (2021).

6. Rentzsch, P., Witten, D., Cooper, G. M., Shendure, J. & Kircher, M. CADD: predicting the deleteriousness of variants throughout the human genome. Nucleic Acids Res. 47, D886–D894 (2019).

7. Forwood, C. et al. Integration of EpiSign, facial phenotyping, and likelihood ratio interpretation of clinical abnormalities in the re-classification of an ARID1B missense variant. Am. J. Med. Genet. C Semin. Med. Genet. (2023) doi:10.1002/ajmg.c.32056.

8. Arlt, A. et al. Next-generation phenotyping in Nigerian children with Cornelia de Lange syndrome. Am. J. Med. Genet. A e63641 (2024).

9. Carrer, A. et al. Application of the Face2Gene tool in an Italian dysmorphological pediatric clinic: Retrospective validation and future perspectives. Am. J. Med. Genet. A (2023) doi:10.1002/ajmg.a.63459.

10. Richards, S. et al. Standards and guidelines for the interpretation of sequence variants: A joint consensus recommendation of the American College of Medical Genetics and Genomics and the Association for Molecular Pathology. Genet. Med. 17, 405–424 (2015).

11. Tavtigian, S. V. et al. Modeling the ACMG/AMP variant classification guidelines as a Bayesian classification framework. Genet. Med. 20, 1054–1060 (2018).

12. Hustinx, A. et al. Improving Deep Facial Phenotyping for Ultra-Rare Disorder Verification Using Model Ensembles. in Proceedings of the IEEE/CVF Winter Conference on Applications of Computer Vision 5018–5028 (2023).

13. Lesmann, H. et al. GestaltMatcher Database - A global reference for facial phenotypic variability in rare human diseases. medRxiv (2024) doi:10.1101/2023.06.06.23290887.

14. Garavelli, L. et al. Neuroimaging findings in Mowat-Wilson syndrome: a study of 54 patients. Genet. Med. 19, 691–700 (2017).

15. Bhasin, M. A. et al. Enhancing Variant Prioritization in VarFish through On-Premise Computational Facial Analysis. Genes 15, 370 (2024).

